# Rationalising neurosurgical head injury referrals: The development and implementation of the Liverpool Head Injury Tomography Score (Liverpool HITS) for mild traumatic brain injury

**DOI:** 10.1101/19004499

**Authors:** Conor SN Gillespie, Christopher M Mcleavy, Abdurrahman I Islim, Sarah Prescott, Catherine J McMahon

## Abstract

**Objectives:** To develop and implement a radiological scoring system to define a ‘surgically significant’ mild Traumatic Brain Injury (TBI), stratify neurosurgical referrals and improve communication between referral centres and neurosurgical units.

**Design:** Retrospective single centre case-control analysis of ten continuous months of mild TBI referrals.

**Setting:** A major tertiary neurosurgery centre in England, UK.

**Participants:** All neurosurgical referrals with a mild TBI (GCS 13-15) during the period of 1^st^ January to 30^th^ October 2017 were eligible for the study. 1248 patients were identified during the study period, with 1144 being included in the final analysis.

**Interventions:** All patients’ CT head results from the referring centres were scored retrospectively using the scoring system and stratified according to their mean score, and if they were accepted for transfer to the neurosurgical centre or managed locally.

**Main outcome measure:** Determine the discriminatory and diagnostic power, sensitivity and specificity of the scoring system for predicting a ‘surgically significant’ mild TBI.

**Results:** Most patients referred were male (59.4%, N=681), with a mean age of 69 years (SD=21.1). Of the referrals to the neurosurgical centre, 17% (n=195) were accepted for transfer and 83% (n=946) were not accepted. The scoring system was 99% sensitive and 51.9% specific for determining a surgically significant TBI. Diagnostic power of the model was fair with an area under the curve of 0.79 (95% CI 0.76 to 0.82). The score identified 495 (52.2%) patients in ten months of referrals that could have been successfully managed locally without neurosurgical referral if the scoring system was correctly used at the time of injury.

**Conclusion:** The Liverpool Head Injury Tomography Score (HITS) score is a CT based scoring system that can be used to define a surgically significant mild TBI. The scoring system can be easily used by multiple healthcare professionals, has high sensitivity, will reduce neurosurgical referrals, and could be incorporated into local, regional and national head injury guidance.

## Introduction

Traumatic brain injury (TBI) is a significant source of morbidity and mortality worldwide, with approximately 69 million affected individuals each year^1^. In England and Wales, 1.4 million patients attend hospital emergency departments following head injuries^1-3^. TBI is an increasingly prevalent problem, with studies reporting a 50%+ increase in emergency department admissions within the last ten years for patients with TBI^4-6^. TBI is also a highly pertinent neurosurgical problem, and accounts for up to 50% of neurosurgical on-call workload^7, 8^. TBI can be classified into mild, moderate, and severe^9, 10^. This classification has been described in a plethora of ways, including the clinical features of the patient (such as post-injury length of amnesia and loss of consciousness), however in the acute phase (first 24 hrs) of a TBI it is recommended to classify it into mild, moderate and severe depending on the presenting Glasgow Coma Scale (GCS), with mild being a GCS of 13-15, moderate being 9-12 and severe being less than 9^11^. Seventy-five to 80% of TBI is classified as mild^12, 13^.

The National Institute for Health and Care Excellence (NICE) guidelines recommend involving neurosurgical centres early if appropriate in TBI management, however the majority of patients referred to neurosurgical centres for management of their mild TBI are not accepted for transfer and are thus managed locally^14, 15^. Indeed, a recent systematic review and meta-analysis suggests that 3.5% of those with a mild TBI go on to receive neurosurgical intervention^15^. In contrast, Neurosurgical referrals for trauma are constantly increasing and have increased by 50% in the last 5 years in the UK^16, 17^. This increase^18^ coincides with recent surveys that suggest referrers find many aspects of referral to a neurosurgical centre difficult, and a lack of training in under- and postgraduate medical programmes indicate that many doctors may not be aware of what injuries in mild TBI constitute a neurosurgical emergency which merits referral^19, 20^. Relative low satisfaction with the referral process and most particularly, the reported unwillingness of on-call neurosurgeons to accept patients to tertiary centres also suggests that there is an issue with communication on both the sides of the referral pathway^21, 22^.

NICE currently recommends TBI patients with a moderate or severe injury to be referred to a neurosurgical unit for advice and input with regards to management and potential transfer. For mild TBI, referral to a neurosurgeon should be considered if an injury is ‘surgically significant’; with those not eligible for this being managed locally according to locally developed pathways^23^.

However, there are currently no national (e.g. NICE) or international guidelines available elucidating the nature of a ‘surgically significant’ mild TBI. This is generally considered to be a mild TBI that warrants referral to an on-call neurosurgeon, and what constitutes this is recommended to be determined by local centres and trust protocols, working together with tertiary neurosurgical centres and emergency departments^23^.

There are several issues with this. Firstly, development of local protocols could lead to variability in referral and acceptance rates across centres which has been explored in previous studies relating to the management of TBI^24, 25^. This is more pertinent for referral centres without readily available access to a neurosurgical centre^26^. In many local trusts, all computed tomography (CT) head scans reported as ‘abnormal’ after a mild TBI are often referred to neurosurgical centres, leading to inappropriate referrals^17, 25, 27^. Indeed, at our centre roughly 82% of these referrals are not accepted for transfer and managed locally. This accounts for up to 1200 extraneous referrals per centre per year^28^. As very few mild TBI patients end up receiving neurosurgical management, this means that most referrals are not accepted for transfer to neurosurgical centres and are managed locally^17, 29^. There is little data available regarding the scope of avoidable neurosurgical referrals for mild TBI and the impact that being able to identify this may have. Given that NICE is responsible for national guidance, a national pathway and method of determining a surgically significant mild TBI would help stratify patients that need to be referred to a neurosurgical centre for advice, management or potential transfer. A recognised guidance and scoring system would also reduce avoidable referrals (patients that would be referred to a neurosurgeon on-call only to be later not accepted) thus reducing time spent awaiting the response to the referral by the local referring hospital and associated healthcare costs. In addition, a method of grading the severity of a mild TBI would provide quantification of the severity of mild TBI between referring hospital and neurosurgical centre, facilitating communication and improving satisfaction rates with the referral process.

Our hypothesis was that by stratifying the number of patients that need to be referred based on referral criteria, we would reduce the number of those avoidable. This would improve workload for on-call neurosurgeons and, if this was a numerically scored system, it would provide an objective and documentable method of communication between the referrer and the neurosurgical centre.

## Methods

### Summary

The authors used advice from senior consultant neurosurgeons with a specialist interest in neurotrauma at the Walton Centre NHS Foundation Trust, a specialist tertiary centre for neurology and neurosurgery, as well as a local research and trauma network that includes radiologists, surgeons of other disciplines, specialist trauma nurses and consultant trauma surgeons to propose and develop the scoring system. This was then correlated with a radiology specialist trainee (the principle developer of the scoring system) to assess feasibility and relatability of the score. This was then fed back to the neurosurgical team for approval.

To detect the margin for the study, we accessed referrals for two months of mild TBI referrals from the ORION online neurosurgical referral system as a pilot study in 2017 to estimate how many referrals were accepted compared to not accepted, and the potential number of referrals that could be managed locally and thus did not need to be referred^28, 30^.

As the vast majority of TBI patients receive a CT scan on arrival to most referral centres prior to neurosurgical referral, it was decided to base the scoring system on the results and injuries accrued from this. This would enable the scoring system to be deciphered and interpreted by all radiologists that report the scan, in addition to specialist doctors such as emergency medicine and neurosurgical trainees and consultants. This would also enable the score to be calculated rapidly.

The scoring system was drafted primarily by the Radiology specialist trainee (CML) and reviewed by consultants at our centre, as well as presented at numerous trauma and departmental meetings to review, discern and make modifications before the study. This was then reviewed by a consultant neurosurgeon with a specialist interest in trauma (CM) and finalised.

### Explanation of scoring parameters

The scoring system contains different categories broadly based on anatomical location of radiologically apparent injuries. To calculate the score, the radiological injuries the patient has sustained from each injury category are combined to form a cumulative total with the following outcomes. Scores of ‘0’, ‘1’ or ‘2’ were defined as a “none surgically significant injuries”- and patients with this score can be managed according to local guidance and protocol at the referral centre (e.g. a hospital emergency department). A score of ‘3’ or more was determined as a surgically significant injury that should prompt a referral to a neurosurgical centre for guidance with regards to management and potential transfer. This may be because of the high mortality and morbidity associated with a single injury (e.g. Extradural haemorrhage (EDH)) or a combination of multiple concerning injuries (e.g. multiple contusions) ^31, 32^. The scoring system and how to interpret it using a clinical example are outlined in Figures 2-3.

**Figure 1.**
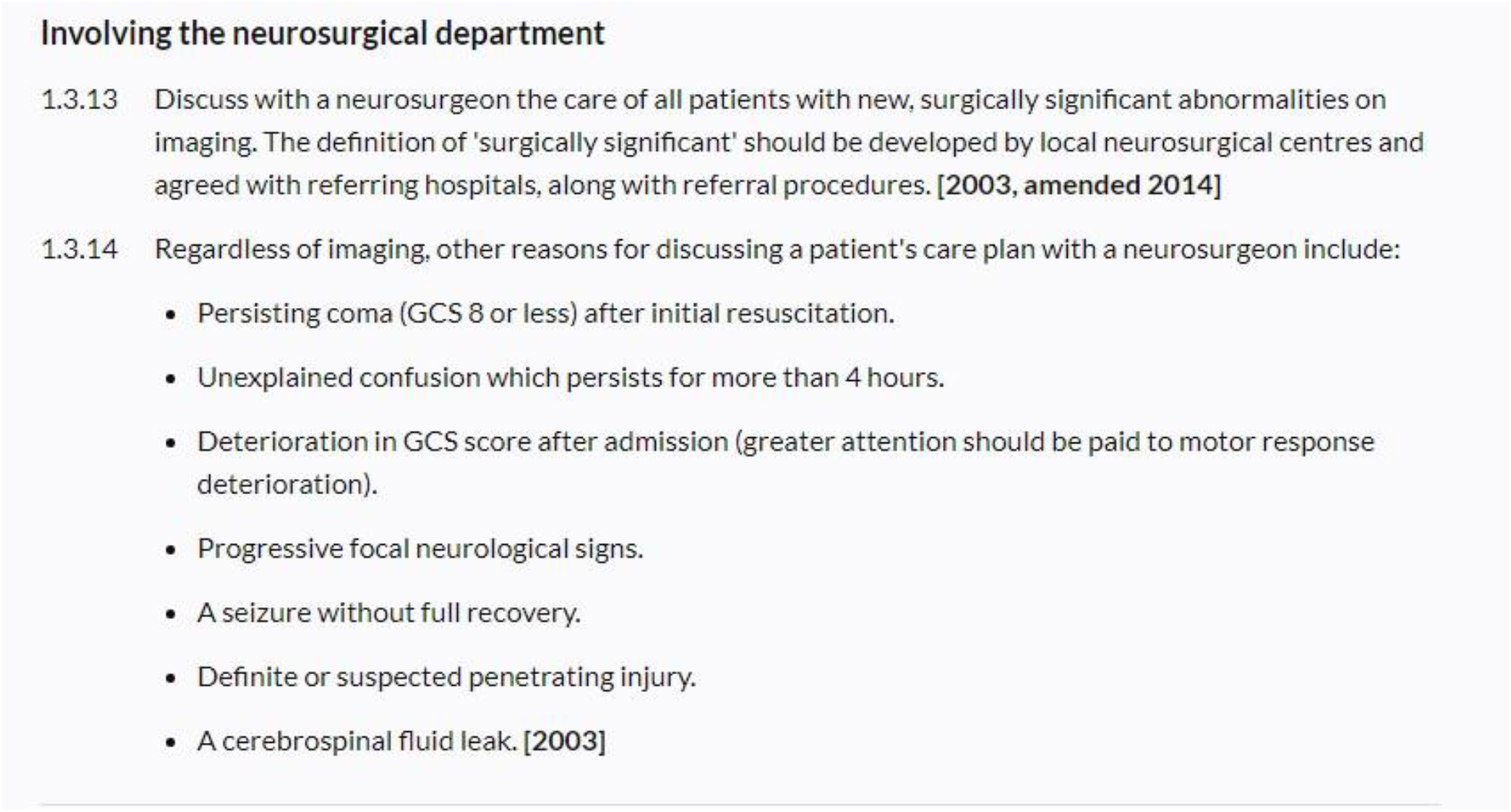
Current NICE head Injury guidance (correct as of July 2019)

**Figure 2.**
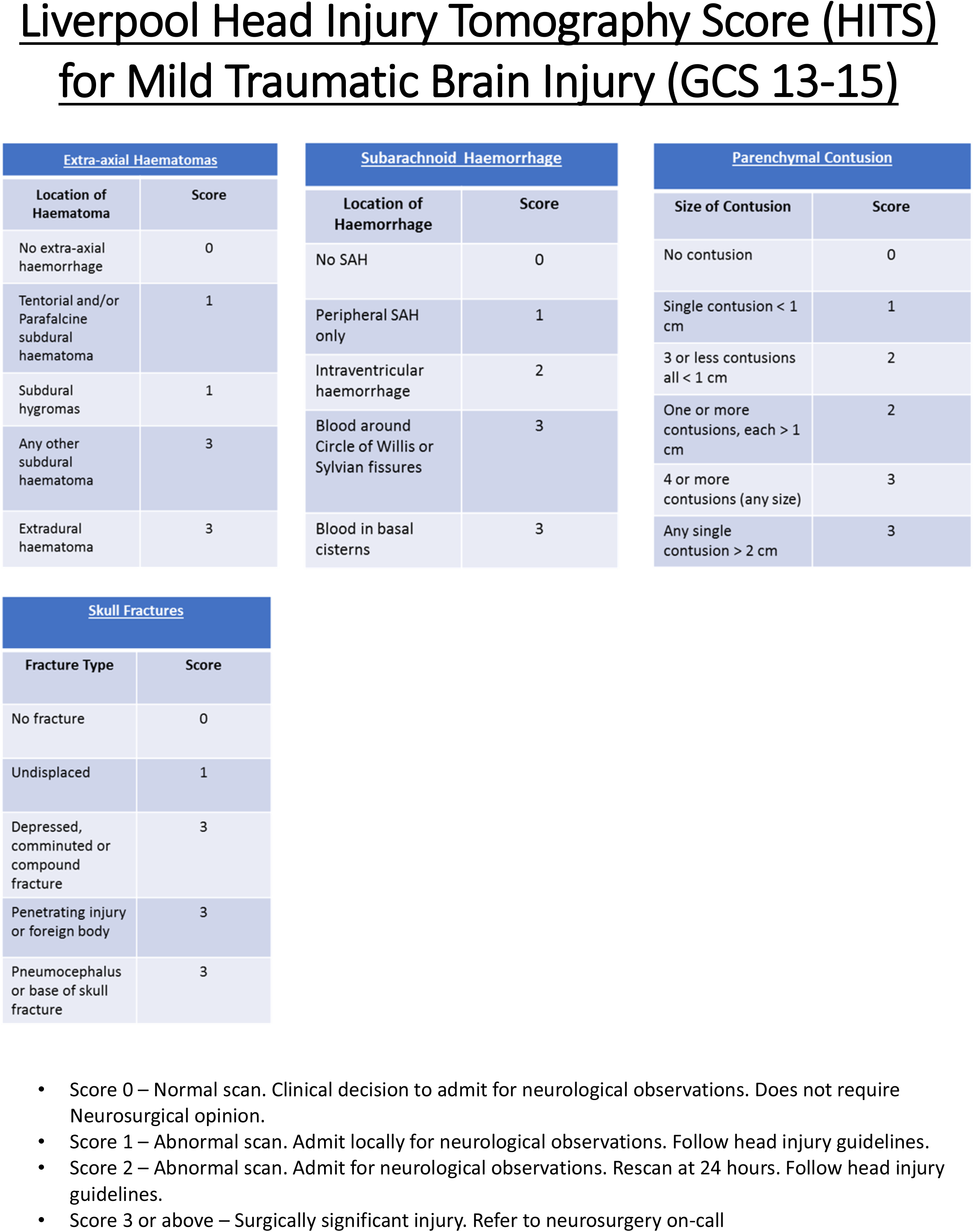
Liverpool Head Injury Tomography Score (HITS) for mild TBI. •Score 0 – Normal scan. Clinical decision to admit for neurological observations. Does not require Neurosurgical opinion. •Score 1 – Abnormal scan. Admit locally for neurological observations. Follow head injury guidelines. •Score 2 – Abnormal scan. Admit for neurological observations. Rescan at 24 hours. Follow head injury guidelines. •Score 3 or above – Surgically significant injury. Refer to neurosurgery on-call

**Figure 3.**
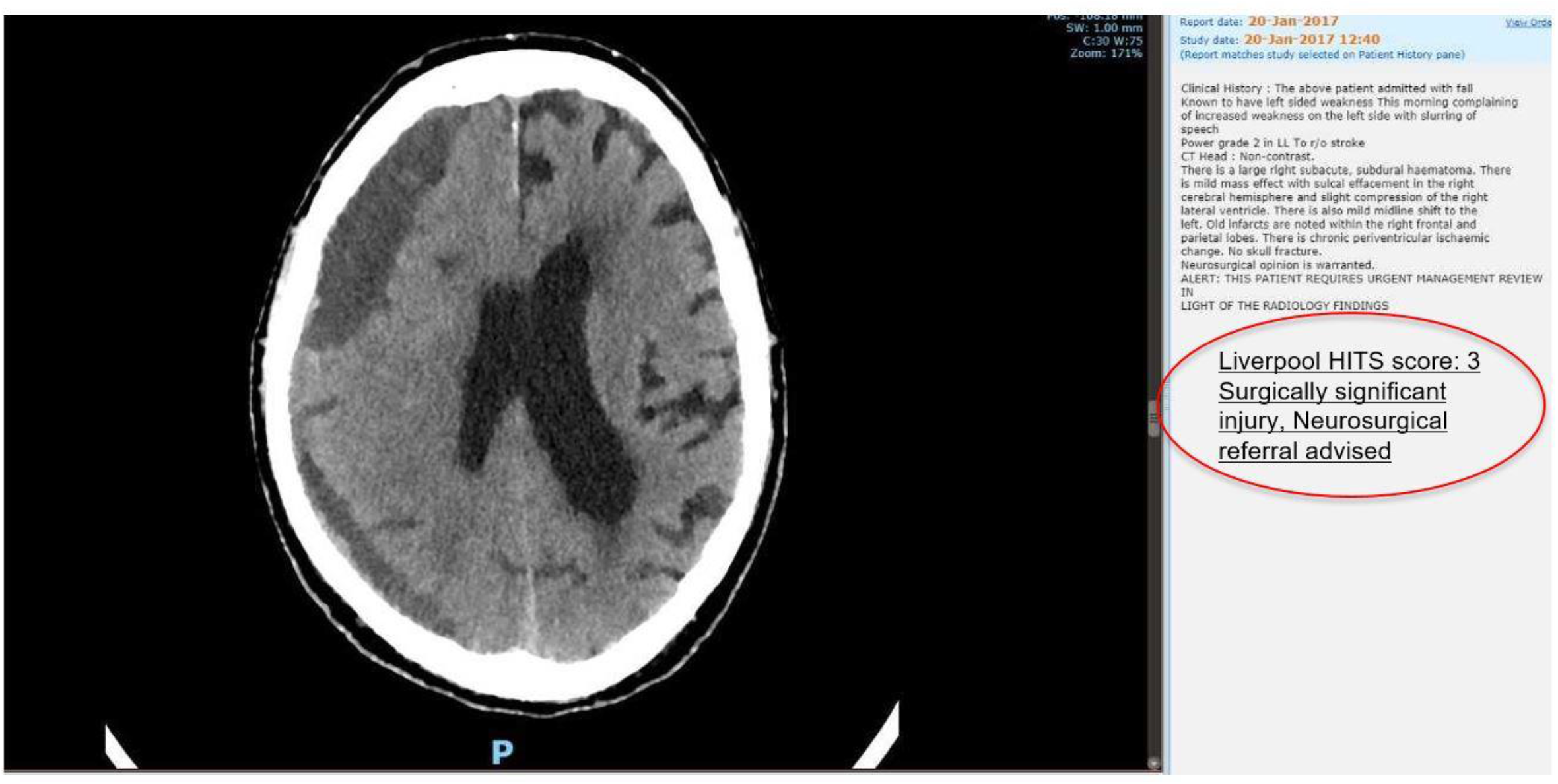
Example case using the Liverpool HIT score (HITS) in a patient presenting to the emergency department with left sided weakness and a GCS of 14.

**Figure 4.**
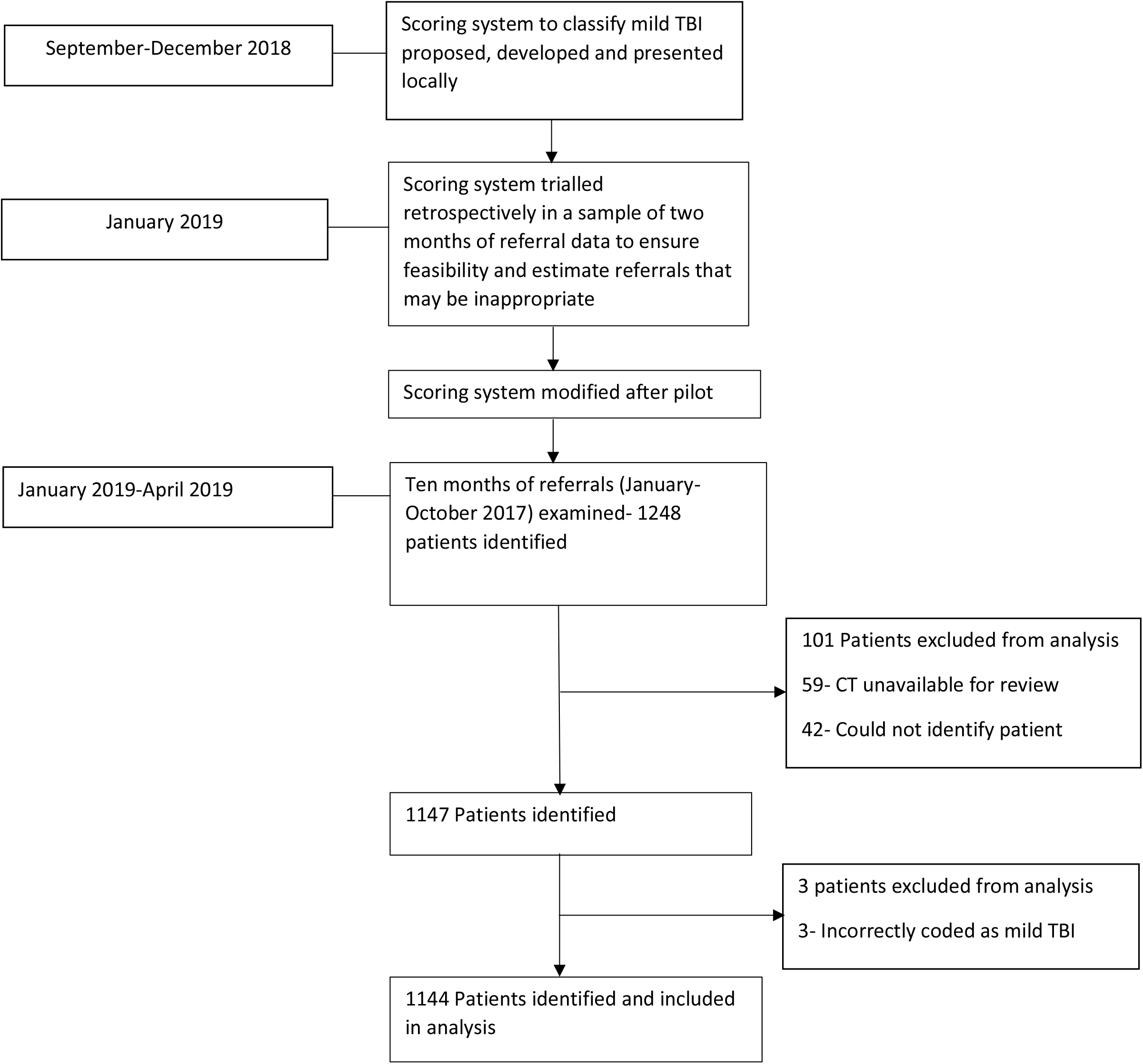
Methods.

**Figure 5.**
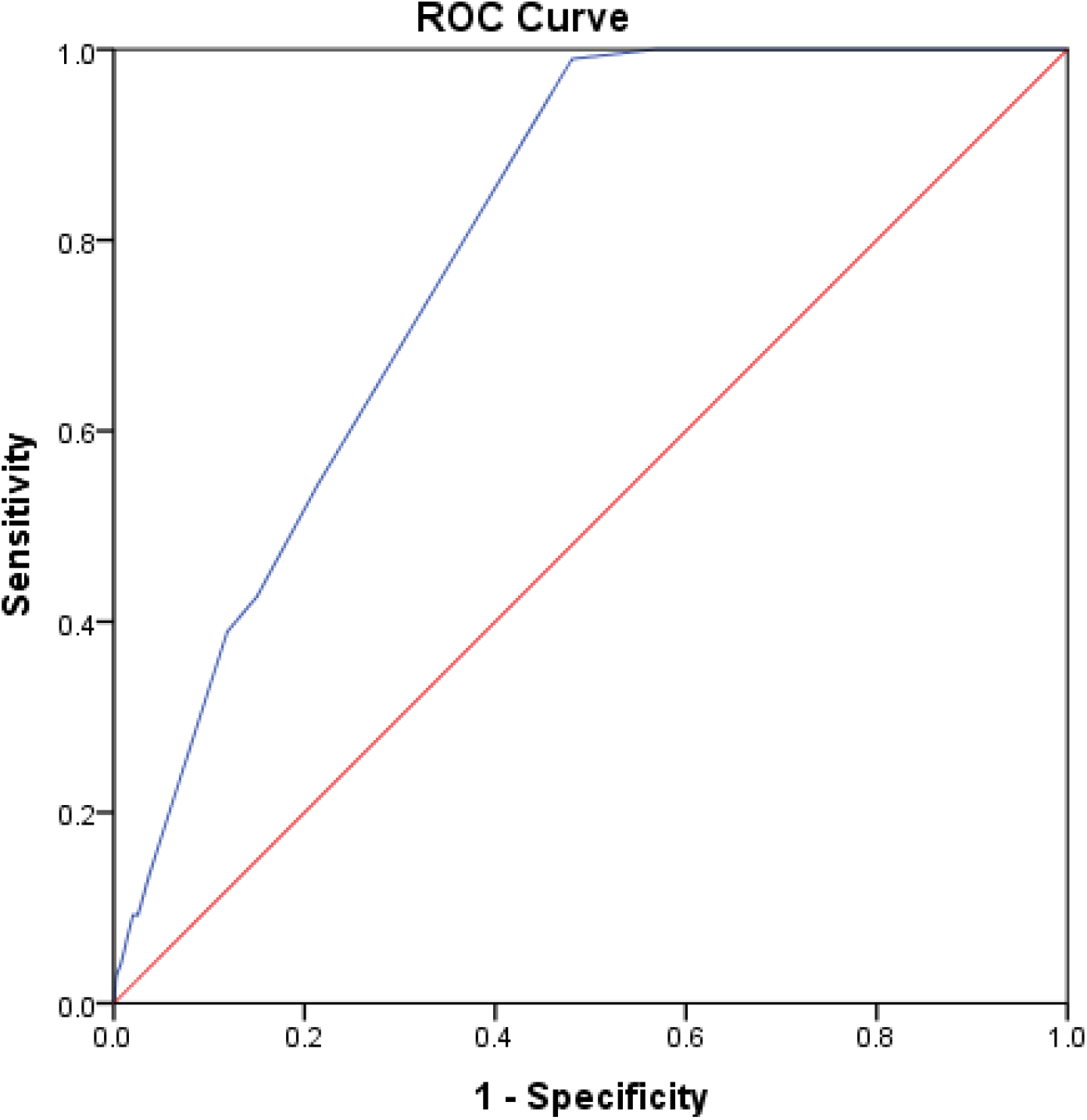
Receiver Operating Characteristic (ROC) curve for Liverpool Head Injury Tomography Score (HITS).

The scoring system was then tested and validated retrospectively. For this we accessed data on every mild TBI referral to our centre for ten continuous months (1^st^ January 2017-30^th^ October 2017). This included every patient that was referred to our centre with a TBI and a GCS of 13-15 during this period. This was accessed with assistance from our local trauma and research network (TARN) and ORION coordinator (SP) to determine if they were accepted for neurosurgical intervention or managed locally. Local audit approval was obtained.

Each individual patient’s referral details and CT head scan (with or without a report from a local radiologist) were accessed. Each individual scan was reviewed and scored retrospectively using the scoring system. This was used to determine how many patients were accepted, the HIT score of each patient and how effective the scoring system was at defining a surgically significant injury. We stratified the patients according to whether they had been accepted or not by our centre for transfer or any form of intervention as our outcome measure. This included non-surgical intervention (e.g. acceptance for monitoring and observation) or surgical intervention (e.g. evacuation of a haemorrhage and intracranial pressure (ICP) monitoring). We then compared the differences between those accepted and those managed locally and their respective HIT scores.

Our second outcome measure was to determine if the score could be safely used prospectively, and that it would not incorrectly categorise a patient as having a ‘non-surgically significant injury’ (i.e. a HIT score of 0, 1 or 2) who was then accepted by the neurosurgical centre for transfer.

To ensure validity and exclude observer bias, after the scores had been calculated a random sample of 50 patients were analysed to check for validity by CML and a consultant radiologist, with no modifications being made to the patients’ original score following this review.

The combined patient details, acceptance status and score were then imported and analysed with descriptive frequencies using SPSS v24.0 (IBM, Armonk, NY, USA). We also carried out sensitivity and specificity analysis, and a Receiver Operating Characteristic (ROC) curve was calculated to determine the overall discriminatory power of the scoring system, with the Area under the curve (AUC) representing the correct prediction rate for the score^33^.

The null hypothesis of the study was that the scoring system would incorrectly categorise patients with a ‘none-surgically significant’ injury and recommend local management, whom ended up being accepted by a neurosurgical centre. If the scoring system were to be effective, it would categorise all patients that were accepted for neurosurgical input as having a surgically significant injury (a score of ‘3’ or more). The number of correct predictions using the score forms the basis of this study, with a diagram of the score development and outline illustrated in Figure 2. The inclusion and exclusion criteria are also outlined below.

### Patient and Public Involvement

Patients or the public were not involved in the design, or conduct, or reporting, or dissemination of our research. As the scoring system and its interpretation, reporting and clinical significance is carried out by healthcare specialists, It was not appropriate to involve patients in the design stage of the study.

## Results

### Identifying number of mild TBI referrals during study period

Of the ten continuous months of referral data examined, 1248 patients were referred to our centre with a suspected mild TBI. Of the 1248 patients eligible for the study, 1144 were included in the final analysis. 101 patients were ineligible for the study, with most of these (n=59) due to inability to access patient CT scans on our referral system. This is due to the neurosurgical referral being from a hospital that is from another county or country, or the referring hospital using an electronic reporting system inaccessible to our centre. In addition, the identity of 42 patients could not be traced and were noted as ‘unknown male’ or ‘unknown female’ on reports and admissions data. These were also excluded.

Finally, further inspection of the data set revealed three patients coded erroneously that were accepted with presenting GCS scores of 6, 7 and 9 respectively and these were excluded due to not qualifying as a mild TBI. This left 1144 patients in the total analysis, who were referred to our centre as a neurosurgical on-call referral, had a mild TBI with a GCS of 13-15, and who could be identified by name and age.

### Patient characteristics and referral outcomes

Males were most likely to be referred due to mild TBI (59.4%, N=681) compared to females (40.6%, N=466). The mean age of referred patients was 69 years (SD=21.1). The most common score overall was 3 points (253 from the not accepted group, 86 from the accepted group). This was due to a large number of subdural haematomas (both acute, chronic and acute-on chronic) being present in the cohort, which scores 3 points (n=283).

Of the 1144 patients referred with a GCS of 13-15 during the ten-month study period, 195 (17.0%) were accepted by the centre for transfer and 949 (83.0%) were not accepted. There was a statistically significant difference (P<0.001) in the mean scores of the accepted group (4.8 points [SD=2.2]) compared to the non-accepted group (2.42 points [SD=2.311]). The range of HIT scores for the accepted group was 13 (2-15), and the range of scores for the local management group was 15 (0-15).

Of those patients not accepted, 454 (47.8%) scored three or more points on the HIT score and thus should have been referred to the neurosurgical centre. However, 495 patients (52.2%) scored 0, 1 or 2 points on the scoring system. They therefore were referrals for patients that could have been managed locally according to local mild TBI protocol without neurosurgical discussion.

Of those accepted, almost all patients (99.0%, n=193/195) had a HIT score of 3 or greater (mean score 4.79). This means that the score would have been correct in establishing surgically significant referral injuries for all but two patients.

Of those accepted, two patients scored less than 3 but were still admitted to the neurosurgical centre. On further investigation, this was because one of the patients also had severe cervical spine injuries that included a fracture to the odontoid peg and was accepted with a view to treat the spinal injuries, and the second patient was accepted due to a lack of beds at our nearby trauma centre and the patient was admitted for neurological observations as a courtesy to the trauma centre.

#### Sensitivity and specificity analysis

The study was 99.0% sensitive including the two aforementioned patients (100% sensitive when excluding them) and 51.9% specific. The positive predictive value (PPV) of the scoring system was 29.8%, and the negative predictive value of the scoring system was 99.6%. The Receiver Operating Characteristic (ROC) curve used to examine sensitivity and specificity together is shown below. Diagnostic power of the model was fair, with an Area under the curve (AUC) of 0.791 with a 95% Confidence Interval (CI) of 0.76 to 0.82^33^.

## Discussion

Perhaps the most notable point of our study was that roughly 83% of referrals to neurosurgical centres for mild TBI are not accepted for transfer, and of these approximately half are likely to not warrant neurosurgical intervention. Thus if using the scoring system these patients would be safely managed locally. This would improve communication between both referring and neurosurgical centres, reduce workload for on call neurosurgeons, and quantify which types of TBI are most likely to require intervention.

From our results for ten months of referral data, implementing the scoring system would reduce mild TBI referrals by up to 600 referrals per neurosurgical centre per year. Based on the sensitivity of the study, almost none of these patients would require a transfer to a neurosurgical centre. However, this figure may vary depending on the size of the centre and acceptance variability.

The sensitivity is very high, which indicates that the scoring system is able to delineate what a ‘surgically significant’ mild TBI is, in addition to reducing the number of inappropriate referrals. This means that the score is highly unlikely to miss any surgically significant injuries, and thus will recommend referral for almost all injuries that go on to be transferred to a neurosurgical centre. The specificity is not high which indicates that the score becomes less accurate for predicting need for admission due to TBI as the mean score increases. Analysing our admissions data, this was mainly due to the presence of patients that were deemed ‘not appropriate for neurosurgical intervention’ whether this be due to catastrophic injury, increasing age or multiple co-morbidities. In this situation, patients would score highly on initial assessment but not be accepted by a centre, creating a false positive result. These patients will therefore often be rejected by the centre for reasons not linked to the severity and score of the mild TBI.

The study included all patients referred to the centre for ten consecutive months, indicating a real time analysis of TBI referrals. In addition, we were able to collect over 1200 patient cases and validate the scoring system using this data. The fact that the score does not miss out any patients or at least very few patients that would have been accepted also validates it as a highly sensitive marker of surgically significant TBI.

Perhaps the second benefit of this scoring system is it appears to be proficient in determining a surgically significant mild TBI-in that we mean a mild TBI that will require transfer to a neurosurgical centre for any reason. Of the 195 patients accepted for transfer, only two had a HIT score of less than the referral cut-off of three, and these were accepted for other reasons outside of injury severity score. The first patient was accepted due to having complex cervical spine injuries and was accepted under the spinal team, and one patient was accepted due to a lack of general trauma beds at a nearby trauma centre.

This is the first scoring system of its kind to the authors’ knowledge. There are CT based scoring systems that accurately predict TBI mortality (such as Marshall and Rotterdam scores)^34-36^ however none of these are specific to mild TBI, nor do they reference referral criteria for being accepted to a neurosurgical unit^37^. This means the potential impact on practice is high, as there is no current way of deciphering a surgically significant mild TBI at the level of national guidance. Moreover, the scoring system is easy to use and can be calculated quickly by healthcare professionals.

It is known that many patients suffer psychological and long-term residual effects after a mild TBI^3, 38, 39^, and thus the scoring system is not intended to discourage head injury referrals to specialist care, rather to stratify patients that require acute neurosurgical referral. Indeed, at our centre we have implemented a mild TBI outpatient clinic to facilitate this and be utilised in conjunction with the scoring system for patients not referred acutely.

### Implications for policy

Our results have several implications for policy. As there is currently no national guidance, and no other UK units currently have such a scoring system in place according to a recent trauma meeting. It may be possible for this score to be utilised on a local, regional and national scale. In addition, the lack of specific national guidance on this topic makes the scoring system particularly pertinent. Additionally, given the previously discussed dissatisfaction with the on-call referral process, this score has a place in placating necessary referrals and improving this process.

The score is currently being implemented prospectively at our centre with a view for other centres to use the score upon finalisation and publication. It is hoped that, as the implementation of the scoring system increases, it will be modified based on evolving feedback to improve the efficacy, awareness and effectiveness of the score.

### Study strengths and limitations

There are several limitations to this study. Firstly, the study was carried out retrospectively at a single tertiary centre with a wide population reach. It is unclear how this may affect the number of referrals and those saved each year, in addition to the mechanism of injury distribution for each centre. However, we analysed continuous data from ten consecutive months of referral data at a UK tertiary centre, making the results pertinent nationally. Second, the retrospective nature of the study does not enable a real-time estimate of the effectiveness of the score to be established; however, the scans were taken at the point of referral to best simulate this enabling translation into real life practice.

Third, the score lacks specificity as a few patients with high scores were not accepted, and this was primarily down to intervention not being plausible or feasible given the patient’s clinical status. Lastly, we were unable to measure patient survival rates for patients who were not accepted for transfer. This is due to differing trust IT systems with the only information available about the patient to us being the referral outcome and does not include management data as this will have been carried out and monitored by the referring centre. Thus, we are unable to completely guarantee that the patients did not receive neurosurgical management elsewhere.

## Conclusions

The Liverpool Head Injury Tomography Score (HITS) is a novel, CT based scoring system to classify mild TBI according to surgical significance and whether the patient requires further neurosurgical centre referral. The scoring system can be used by a myriad of healthcare specialists, is simple to use and has close to 99% sensitivity for predicting a surgically significant mild TBI. This may also be used to stratify and reduce inappropriate referrals to neurosurgical centres by up to 50%, saving workload and improving communication between neurosurgical and referring centres. If implemented correctly, the score could be incorporated into local, regional and potentially national guidance.

**What is known about this topic**

- Traumatic Brain Injury (TBI) is a highly prevalent neurosurgical problem and the number of referrals are increasing year on year
- 75-80% of TBI is classified as mild (GCS 13-15)
- The majority of referrals to neurosurgical centres for patients with mild TBI are not accepted for transfer
- At present there is no national guidance to stratify what mild TBI can be managed locally and what needs to be referred to a neurosurgical centre

**What this study adds**

- The Liverpool head injury tomography score (HITS) is a novel, CT head based scoring system to quantify the severity of mild TBI
- The scoring system is 99% sensitive for detecting a surgically significant mild TBI
- This could be used to stratify patients and reduce avoidable neurosurgical referrals at local, regional and potentially national level.

## Presentations

This work has been presented as an oral presentation by CML at 2019 British Society Of Emergency Radiologists (BSER) 2019 meeting and as an oral presentation by CG at the Society of British Neurosurgeons (SBNS) spring meeting 2019.

## Data Availability

Anonymised statistical data and analysis are available from the corresponding author.

## Author contributions

Conception and design: McMahon, Mcleavy, Gillespie. Acquisition of data: Prescott, Gillespie. Analysis and Interpretation of data: Gillespie, Mcleavy, Islim. Drafting the article: Gillespie, Islim, Mcleavy. Critically revising the article: Gillespie, Mcleavy, Islim, McMahon. All authors agree with the final version of the submitted manuscript.

CG is the lead author and guarantor.

## Ethical approval

Ethical approval was granted by the Walton Centre NHS Foundation trust research and audit team upon commencement of the study.

## Transparency declaration

The lead author (CG) affirms that this manuscript is an honest, accurate, and transparent account of the study being reported; that no important aspects of the study have been omitted; and that any discrepancies from the study as planned have been explained.

## Data sharing

Anonymised statistical data and analysis are available from the corresponding author.

## Copyright

The Corresponding Author has the right to grant on behalf of all authors and does grant on behalf of all authors, a worldwide licence to the Publishers and its licensees in perpetuity, in all forms, formats and media (whether known now or created in the future), to i) publish, reproduce, distribute, display and store the Contribution, ii) translate the Contribution into other languages, create adaptations, reprints, include within collections and create summaries, extracts and/or, abstracts of the Contribution, iii) create any other derivative work(s) based on the Contribution, iv) to exploit all subsidiary rights in the Contribution, v) the inclusion of electronic links from the Contribution to third party material where-ever it may be located; and, vi) licence any third party to do any or all of the above.

## Dissemination declaration

Dissemination of the results to study participants and patient organisations outside of the main study findings is not possible at this stage.

## Preprint declaration

This manuscript is a preprint and should not be disseminated or used to guide policy until formal publication.

## Appendix- Data Tables

**Table 1:** Mean Liverpool HITS scores of referred patients.

| Admitted to Neurosurgery |  |  |  |
| --- | --- | --- | --- |
| Centre | Mean | N | Std. Deviation |
| No | 2.42 | 949 | 2.177 |
| Yes | 4.79 | 195 | 2.311 |
| Total | 2.82 | 1144 | 2.373 |

**Table 2:**
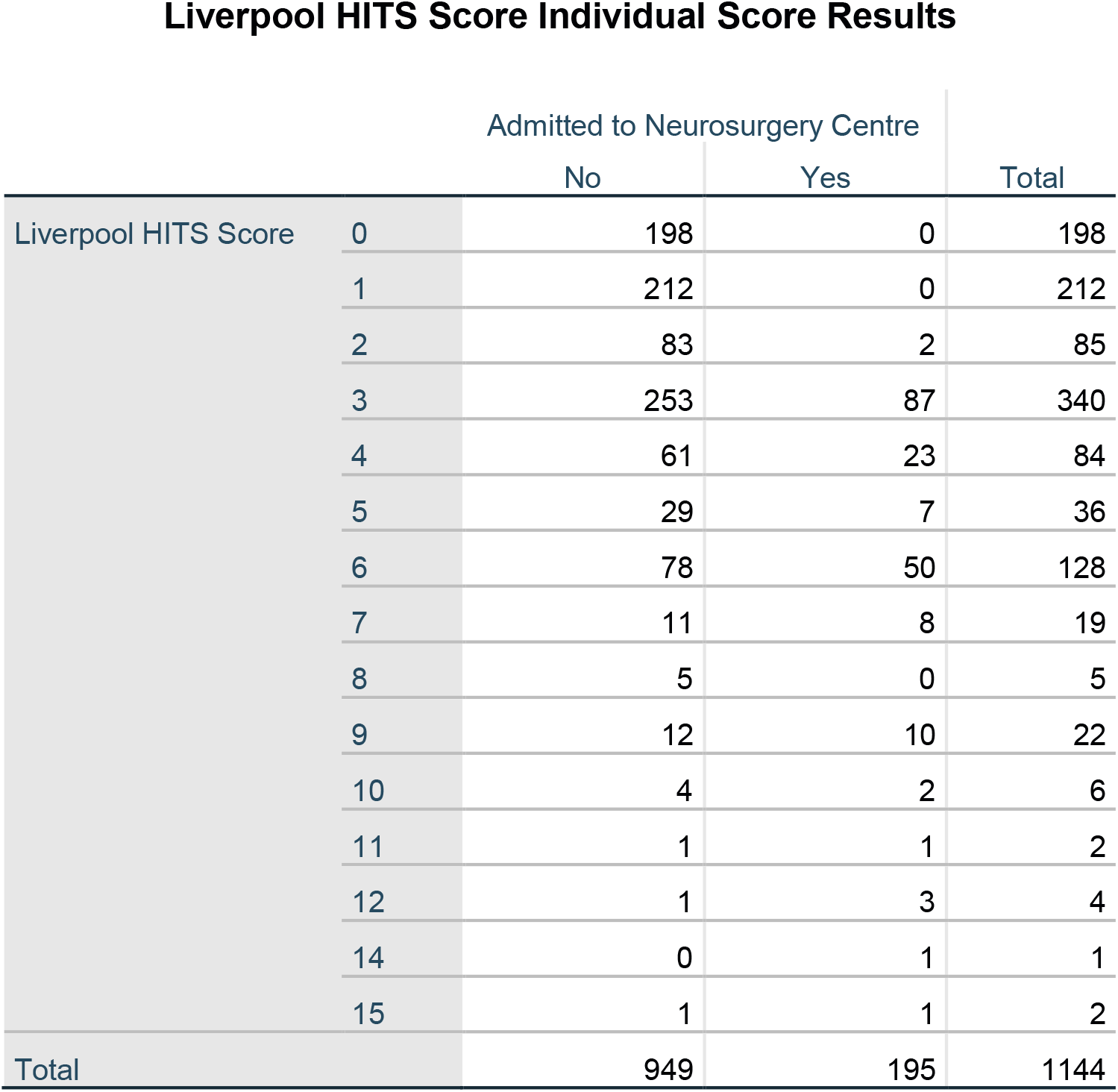
Liverpool HITS Score individual patient score results compared to decision to admit to the Neurosurgical centre

|  |  | Admitted to Neurosurgery Centre |  | Total |
| --- | --- | --- | --- | --- |
|  |  | No | Yes |  |
| Liverpool HITS Score | 0 | 198 | 0 | 198 |
|  | 1 | 212 | 0 | 212 |
|  | 2 | 83 | 2 | 85 |
|  | 3 | 253 | 87 | 340 |
|  | 4 | 61 | 23 | 84 |
|  | 5 | 29 | 7 | 36 |
|  | 6 | 78 | 50 | 128 |
|  | 7 | 11 | 8 | 19 |
|  | 8 | 5 | 0 | 5 |
|  | 9 | 12 | 10 | 22 |
|  | 10 | 4 | 2 | 6 |
|  | 11 | 1 | 1 | 2 |
|  | 12 | 1 | 3 | 4 |
|  | 14 | 0 | 1 | 1 |
|  | 15 | 1 | 1 | 2 |
| Total |  | 949 | 195 | 1144 |

